## Supplemental Data 1 for "EXploring the journeys of Patients who End their Calls prior to Triage by NHS111: The EXPECT study"

**Appendix 1: Modification to the original O'Keeffe et al non-avoidable Emergency Department admission criteria**

| **Disposition code** | **Potentially avoidable** |
|---|---|
| ED Treatment complete | Yes |
| Admitted as inpatient | No |
| Streamed to GP / primary care | Yes |
| Left after assessment before treatment | Yes |
| Left before initial assessment | Yes |
| Left after assessment other | Yes |
| Streamed to Urgent Care Centre | Yes |
| Streamed to ophthalmology service | Yes |
| Died in the Emergency Care facility | No |
| Streamed to Amb Care service | No |
| Streamed to mental health service | Yes |
| Streamed to Emergency Department | No |
| Streamed to dental service | Yes |
| Dead on Arrival | No |
| Discharged with Consent | Yes |
| Streamed to pharmacy service | Yes |
| Streamed to falls service | No |
| Streamed to frailty service | No |
