## Appendix 2-Summary of final model for the EXPECT study for "EXploring the journeys of Patients who End their Calls prior to Triage by NHS111: The EXPECT study"

### Kaplan-Meier plots

Simple model looking at cohort only

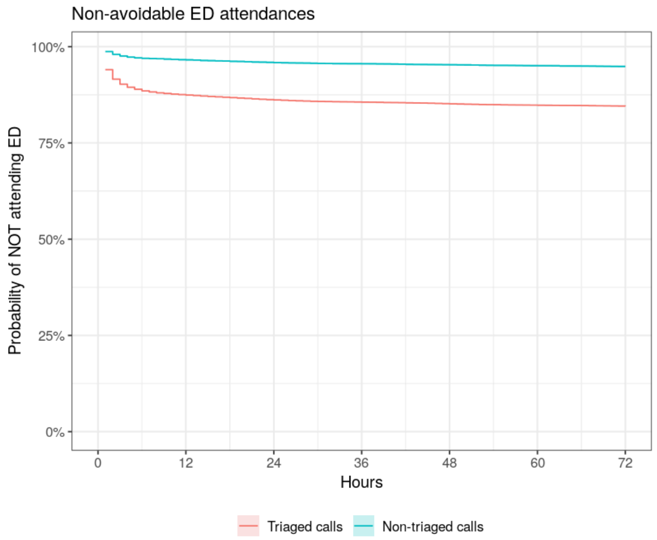


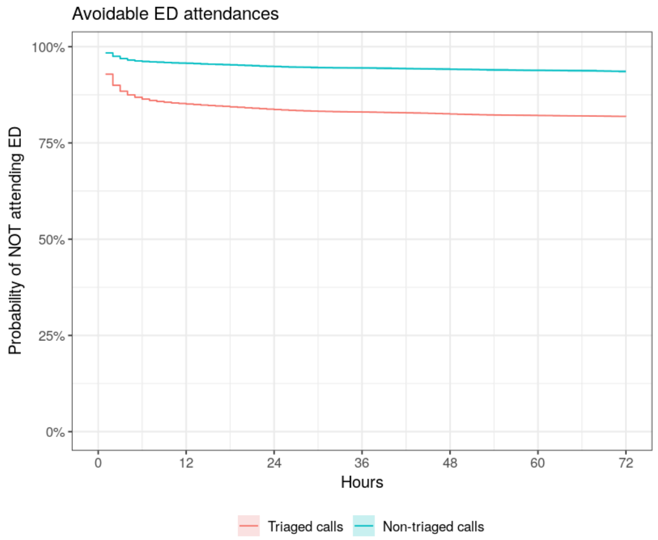


#### Log-log plot

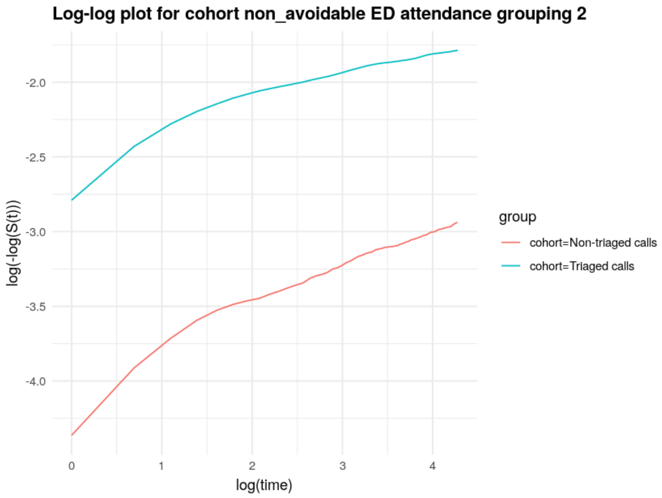


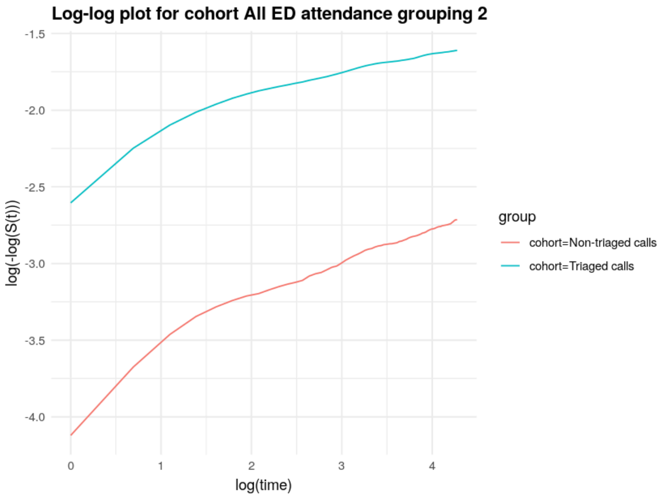


### Cox regression

#### Non-avoidable ED attendances

A tibble: 12 × 8

| term | estimate | std.error | robust.se | statistic | p.value | conf.low | conf.high |
|---|---|---|---|---|---|---|---|
| <chr> | <dbl> | <dbl> | <dbl> | <dbl> | <dbl> | <dbl> | <dbl> |

| term | estimate | std.error | robust.se | statistic | p.value | conf.low | conf.high |
|---|---|---|---|---|---|---|---|
| <chr> | <dbl> | <dbl> | <dbl> | <dbl> | <dbl> | <dbl> | <dbl> |
| cohortNon-triaged calls | 0.32 | 0.03 | 0.03 | -33.29 | 0.00 | 0.30 | 0.34 |
| age | 1.01 | 0.00 | 0.00 | 41.59 | 0.00 | 1.01 | 1.01 |
| sexmale | 0.93 | 0.01 | 0.02 | -5.03 | 0.00 | 0.90 | 0.95 |
| imd_quintile2 | 0.96 | 0.02 | 0.02 | -2.23 | 0.03 | 0.92 | 0.99 |
| imd_quintile3 | 0.90 | 0.02 | 0.02 | -4.51 | 0.00 | 0.86 | 0.94 |
| imd_quintile4 | 0.86 | 0.02 | 0.03 | -5.79 | 0.00 | 0.82 | 0.90 |
| imd_quintile5 | 0.75 | 0.03 | 0.03 | -9.80 | 0.00 | 0.70 | 0.79 |
| ethnicity_simpleAsian or Asian British | 0.98 | 0.02 | 0.02 | -0.88 | 0.38 | 0.95 | 1.02 |
| ethnicity_simpleBlack or African or Caribbean or Black British | 0.81 | 0.06 | 0.06 | -3.34 | 0.00 | 0.72 | 0.92 |
| ethnicity_simpleMixed multiple ethnic groups | 0.96 | 0.06 | 0.11 | -0.42 | 0.67 | 0.77 | 1.18 |
| ethnicity_simpleOther ethnic group | 0.79 | 0.06 | 0.07 | -3.37 | 0.00 | 0.69 | 0.91 |
| ethnicity_simpleUnknown/Refuse to say | 0.86 | 0.02 | 0.02 | -8.02 | 0.00 | 0.83 | 0.89 |

#### All ED attendances

A tibble: 12 × 8

| term | estimate | std.error | robust.se | statistic | p.value | conf.low | conf.high |
|---|---|---|---|---|---|---|---|
| <chr> | <dbl> | <dbl> | <dbl> | <dbl> | <dbl> | <dbl> | <dbl> |
| cohortNon-triaged calls | 0.33 | 0.03 | 0.03 | -35.87 | 0.00 | 0.31 | 0.35 |
| age | 1.01 | 0.00 | 0.00 | 32.92 | 0.00 | 1.01 | 1.01 |
| sexmale | 0.90 | 0.01 | 0.01 | -7.66 | 0.00 | 0.87 | 0.92 |
| imd_quintile2 | 0.96 | 0.02 | 0.02 | -2.48 | 0.01 | 0.92 | 0.99 |
| imd_quintile3 | 0.89 | 0.02 | 0.02 | -5.00 | 0.00 | 0.85 | 0.93 |
| imd_quintile4 | 0.88 | 0.02 | 0.03 | -4.95 | 0.00 | 0.84 | 0.93 |
| imd_quintile5 | 0.77 | 0.02 | 0.03 | -9.44 | 0.00 | 0.73 | 0.81 |
| ethnicity_simpleAsian or Asian British | 1.02 | 0.02 | 0.02 | 0.90 | 0.37 | 0.98 | 1.05 |
| ethnicity_simpleBlack or African or Caribbean or Black British | 0.80 | 0.05 | 0.06 | -3.69 | 0.00 | 0.72 | 0.90 |
| ethnicity_simpleMixed multiple ethnic groups | 0.96 | 0.05 | 0.09 | -0.42 | 0.67 | 0.80 | 1.16 |
| ethnicity_simpleOther ethnic group | 0.87 | 0.05 | 0.06 | -2.16 | 0.03 | 0.77 | 0.99 |
| ethnicity_simpleUnknown/Refuse to say | 0.85 | 0.01 | 0.02 | -9.29 | 0.00 | 0.82 | 0.88 |

### Assessing Goodness-of-Fit using residuals

#### Martingale residuals

##### Non-avoidable ED attendance

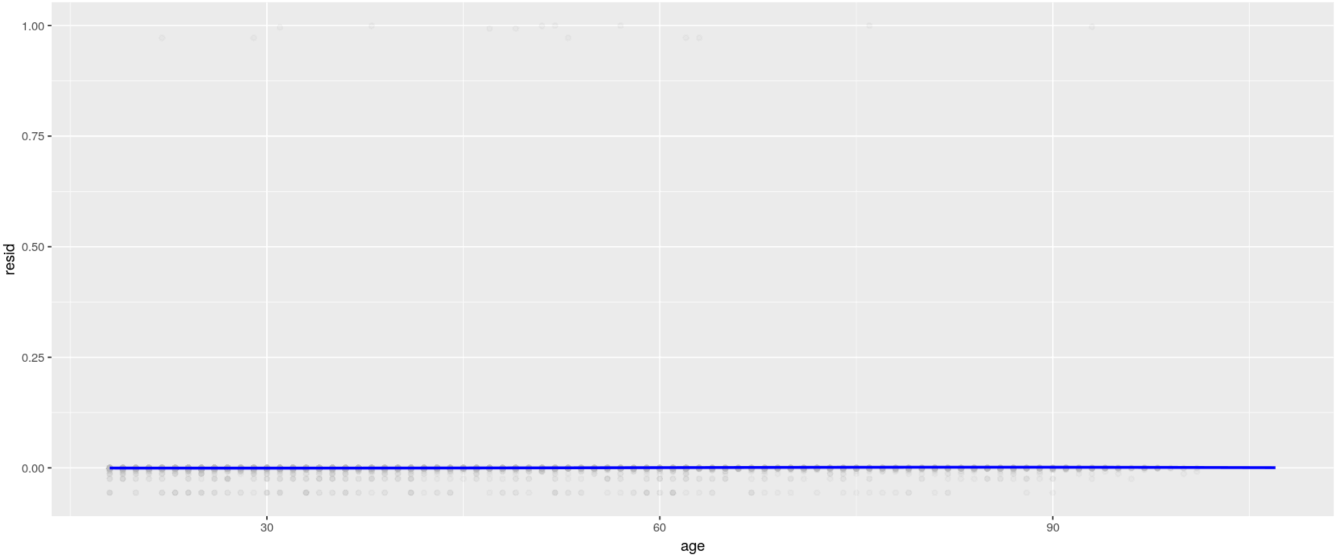


##### All ED attendances

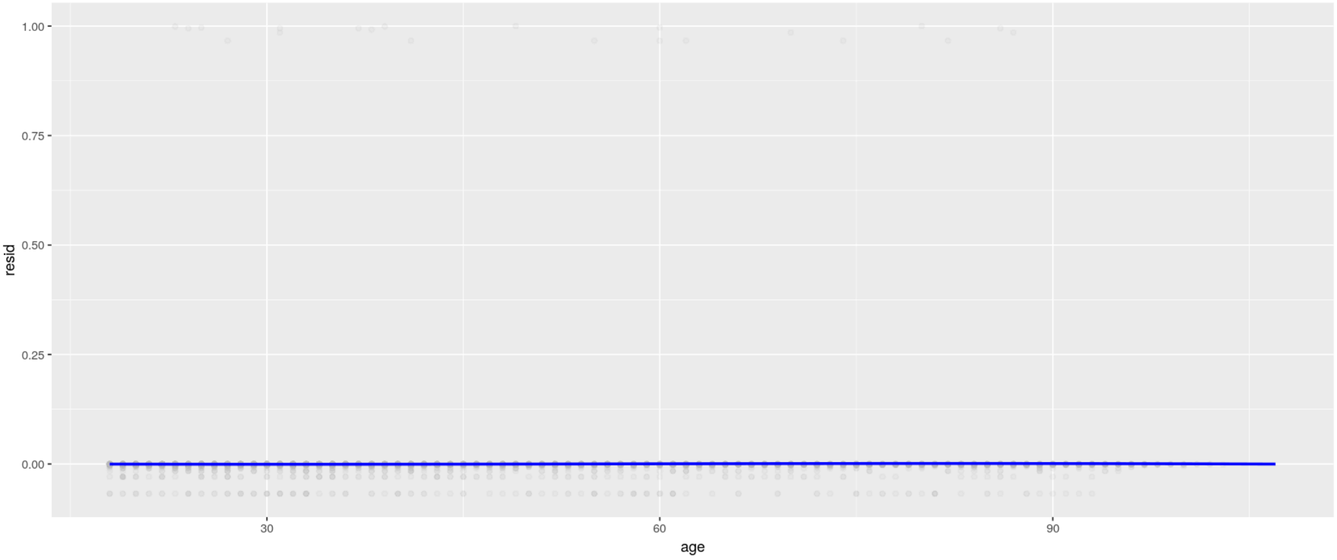


### Checking the proportional hazards assumption

#### Log-log plots

##### Non-avoidable ED attendances

##### Log-log plot for cohort non_avoidable ED attendance

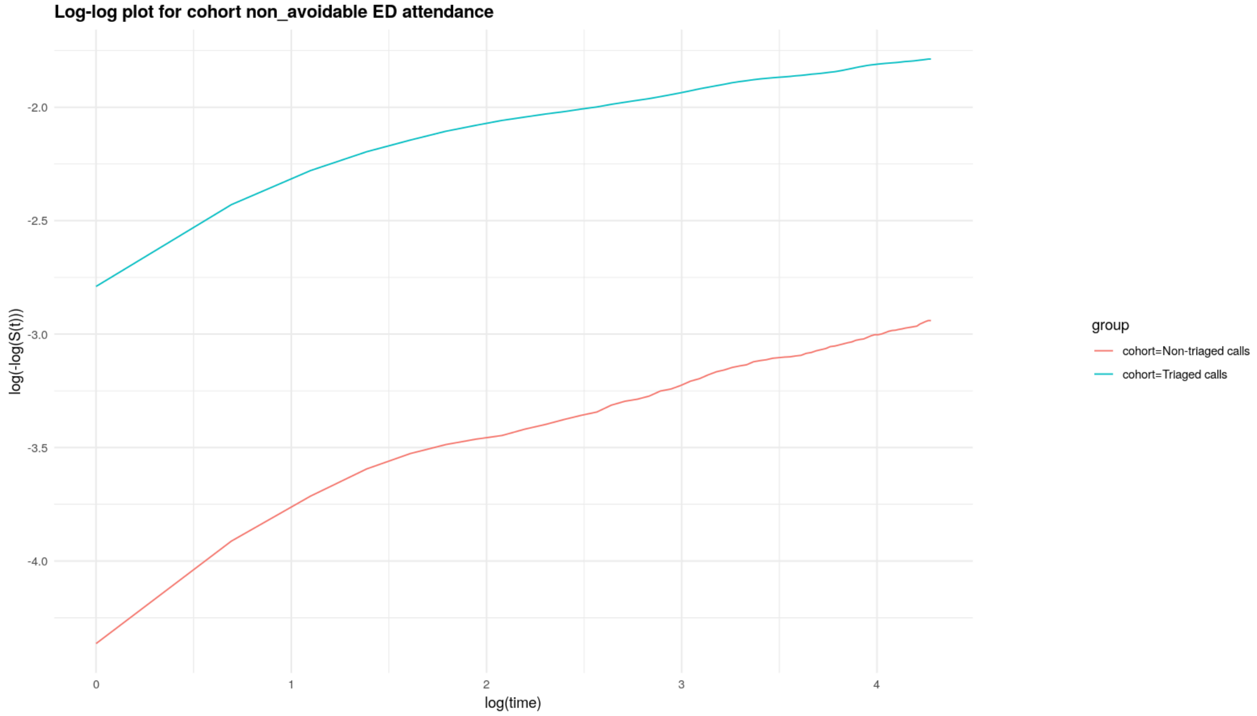


##### Log-log plot for sex non_avoidable ED attendance

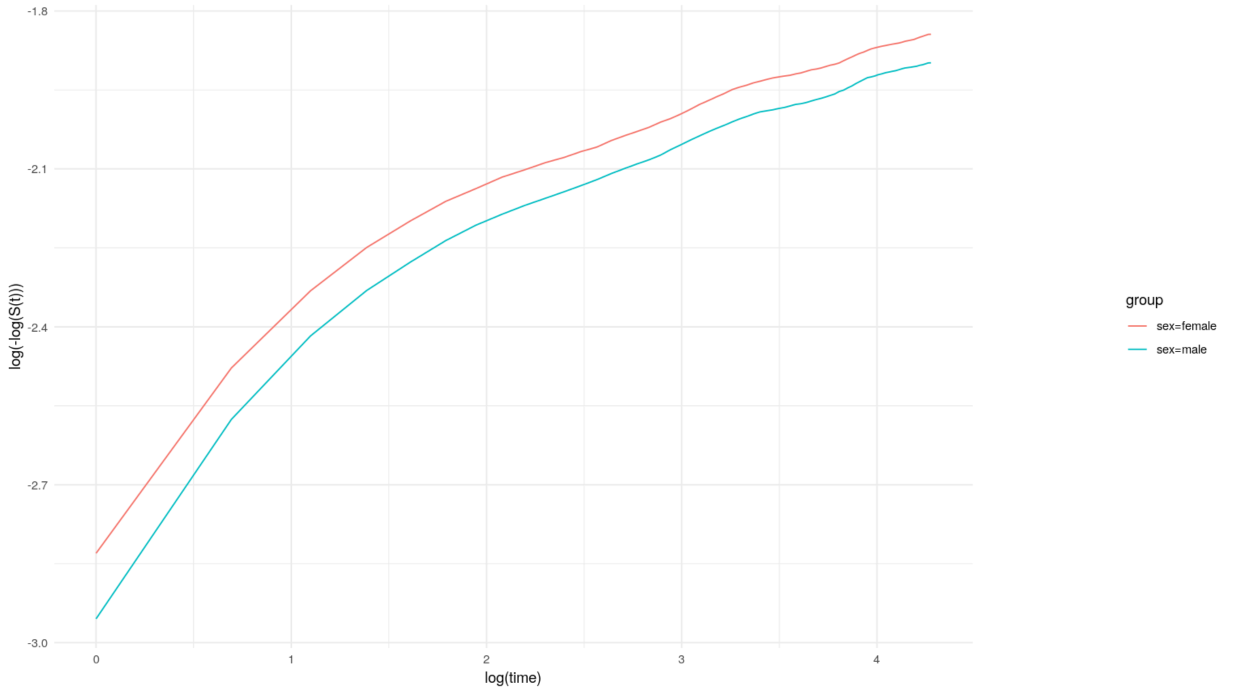


##### Log-log plot for ooh non_avoidable ED attendance

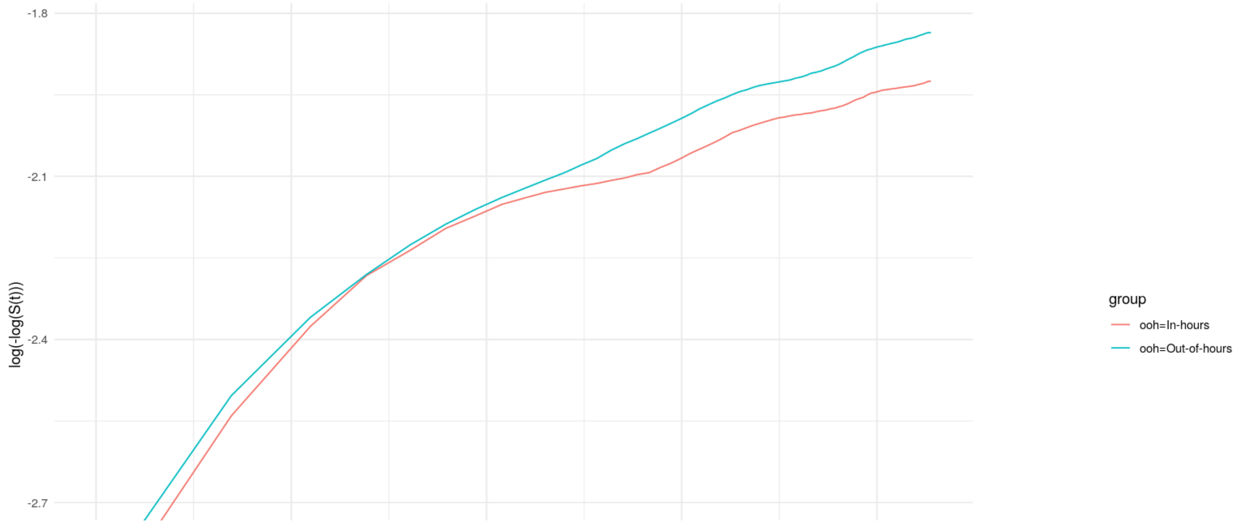

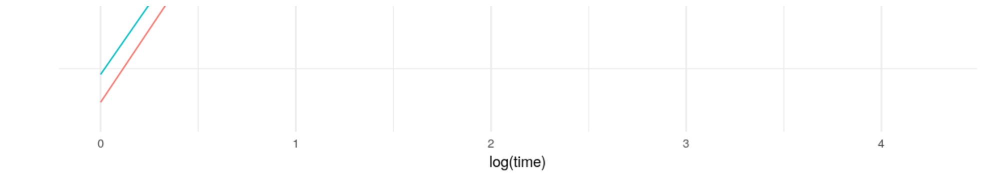


Log-log plot for imd_quintile non_avoidable ED attendance

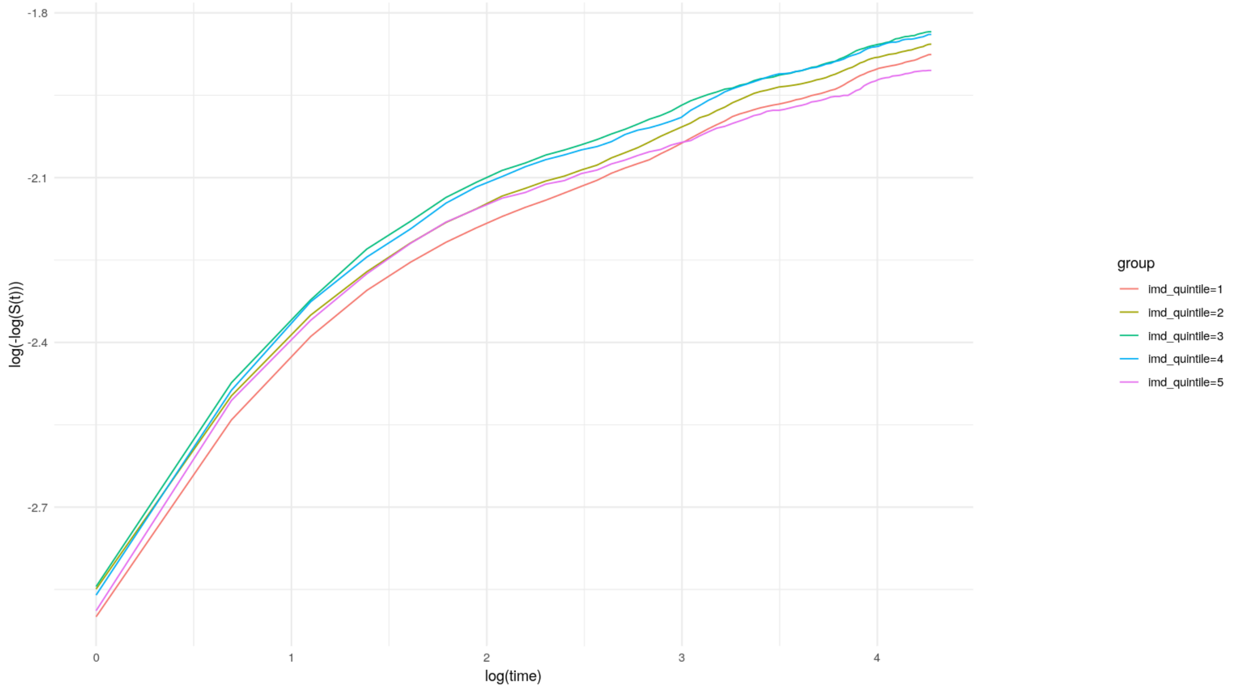


Log-log plot for ethnicity_simple non_avoidable ED attendance

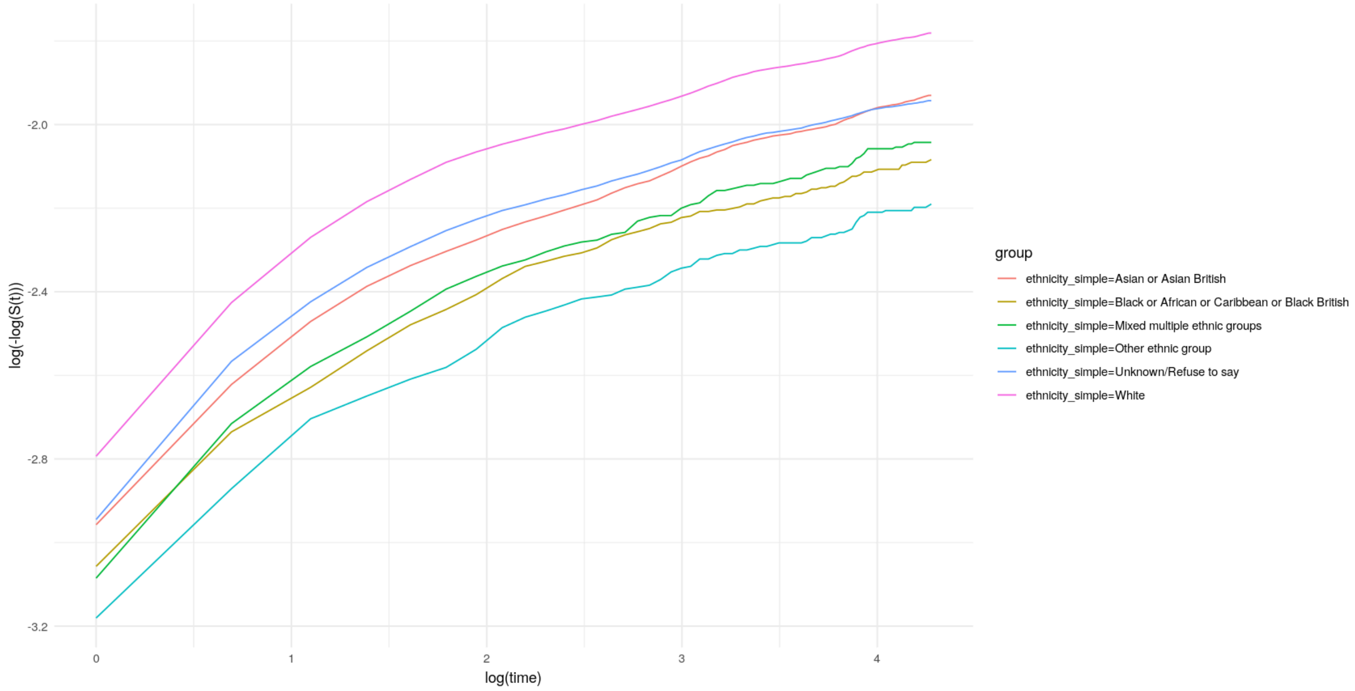


### All ED attendances

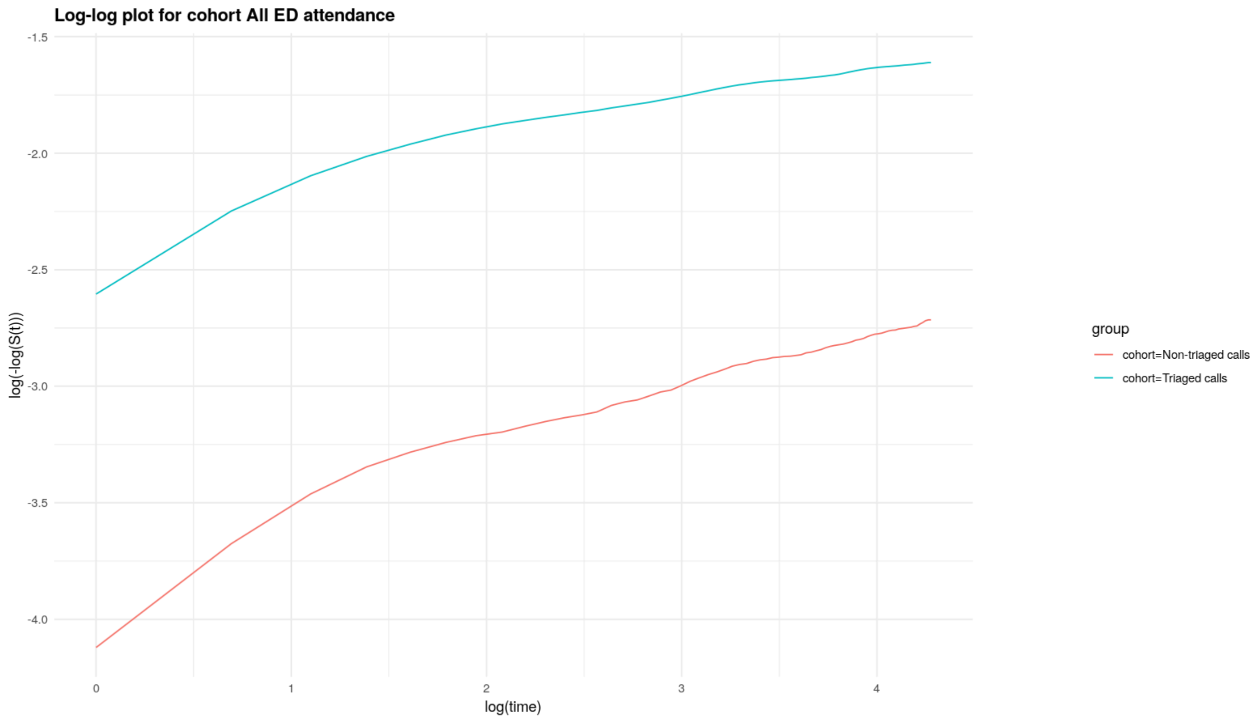

Log-log plot for cohort All ED attendance
log(-log(S(t)))
log(time)
group
cohort=Non-triaged calls
cohort=Triaged calls


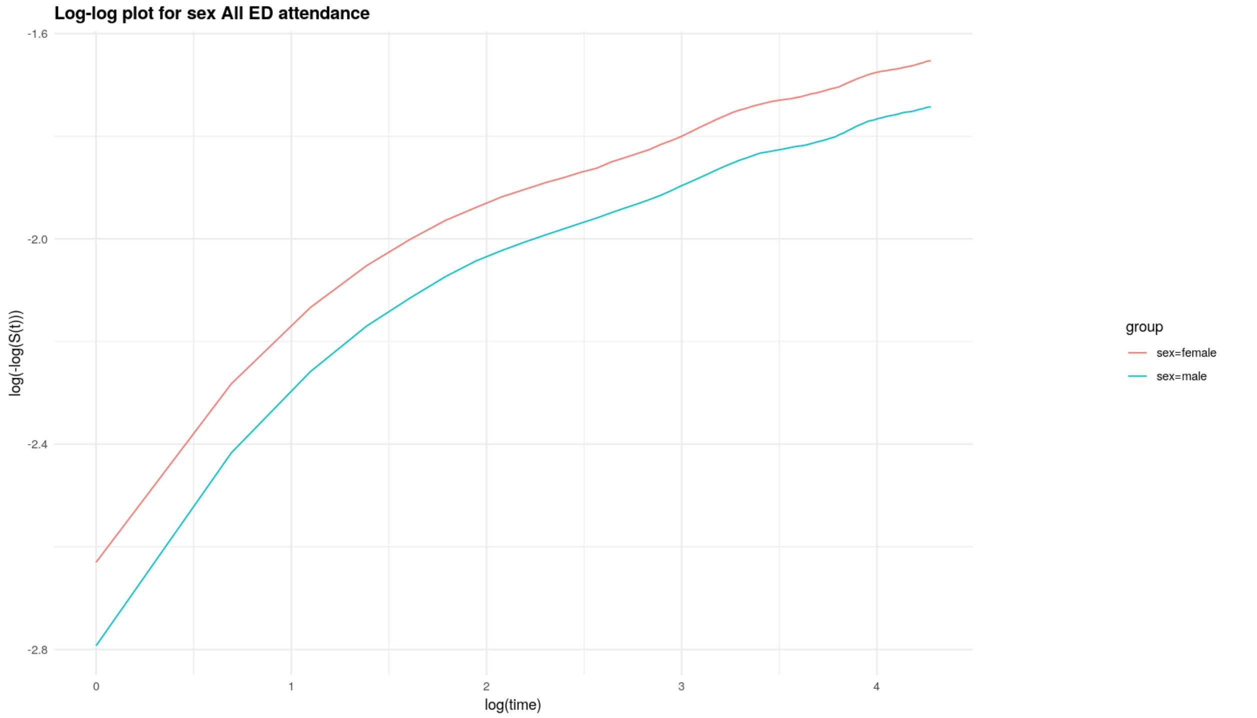

Log-log plot for sex All ED attendance
log(-log(S(t)))
log(time)
group
sex=female
sex=male


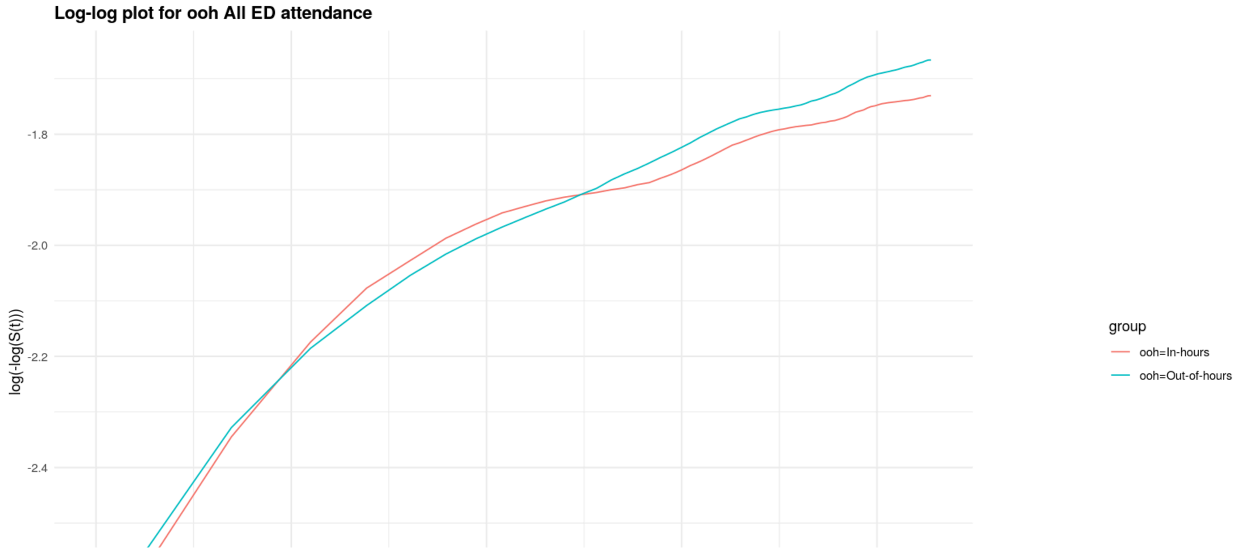

Log-log plot for ooh All ED attendance
log(-log(S(t)))
group
ooh=In-hours
ooh=Out-of-hours

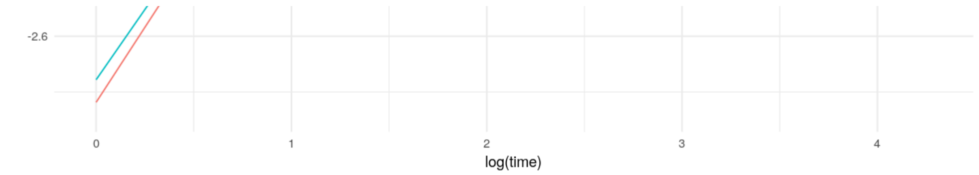


**Log-log plot for imd_quintile All ED attendance**

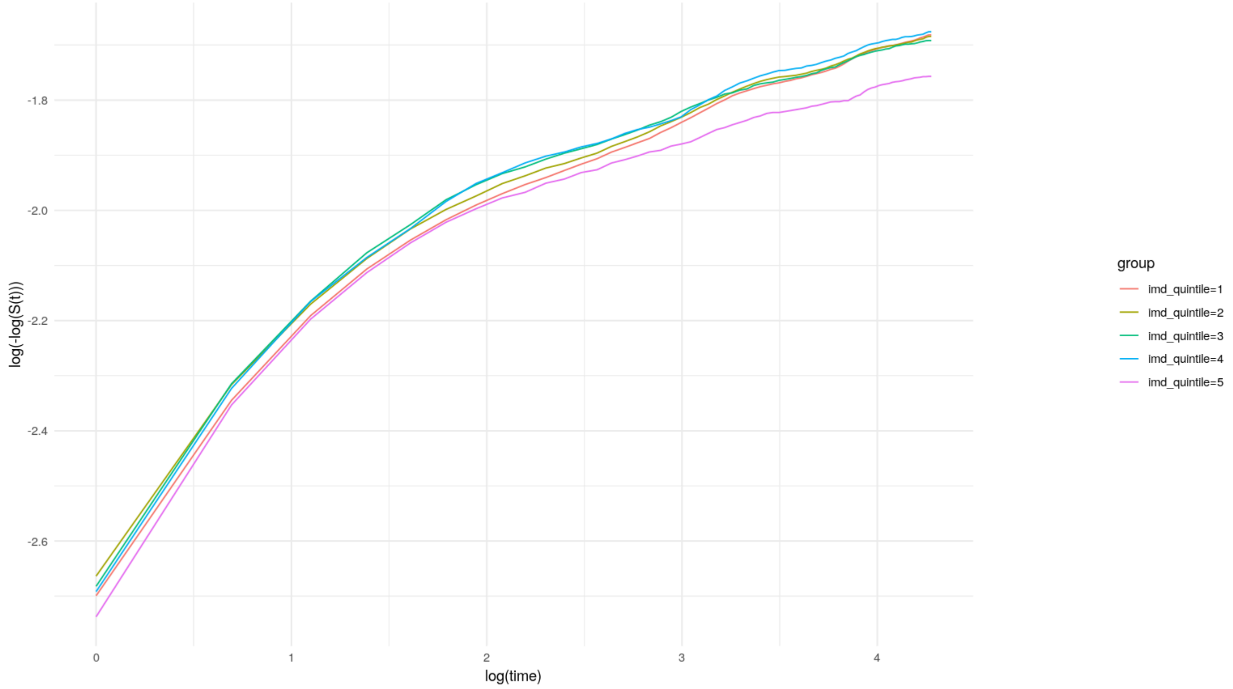


**Log-log plot for ethnicity_simple All ED attendance**

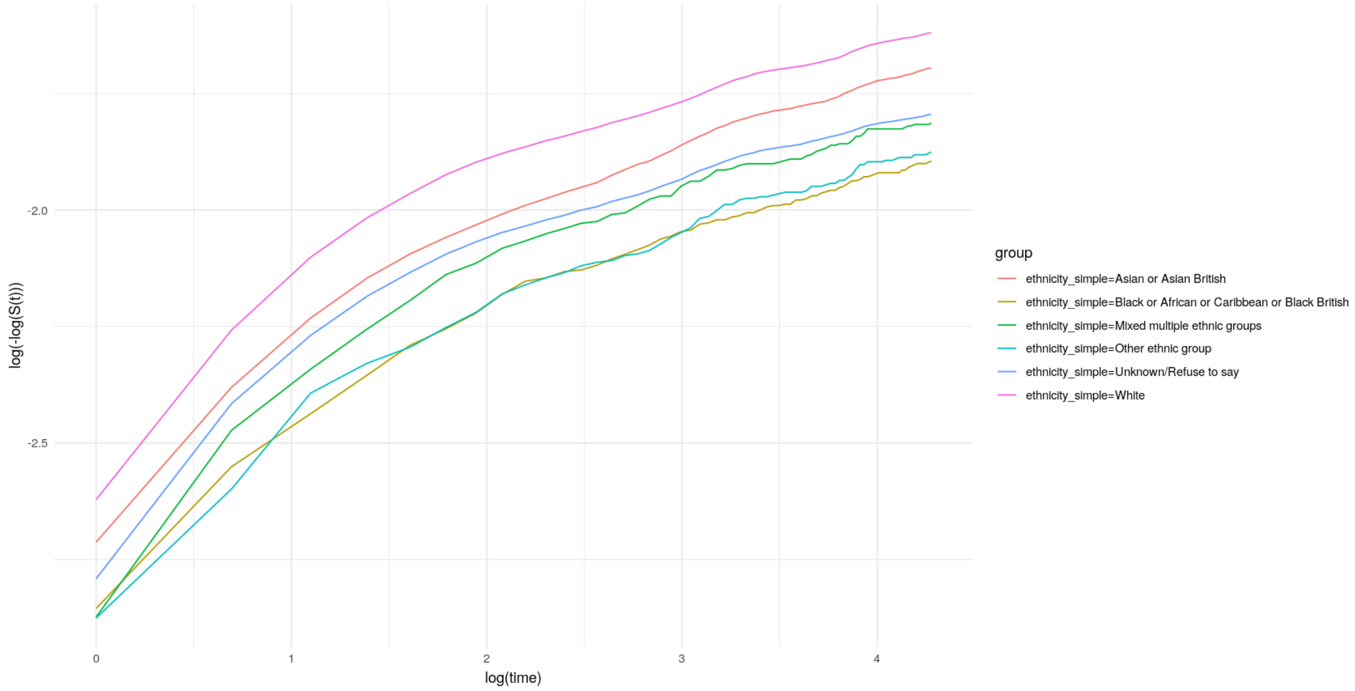


### Non-avoidable ED attendances

```
                 chisq df       p
cohort          105.27  1 < 2e-16
age               3.58  1   0.058
sex              17.15  1 3.5e-05
imd_quintile     11.18  4   0.025
```

```
ethnicity_simple  12.20  5   0.032
GLOBAL           141.11 12 < 2e-16
```

#### All ED attendances

```
                   chisq df      p
cohort           125.059  1 <2e-16
age                0.529  1 0.4670
sex               21.805  1  3e-06
imd_quintile      17.216  4 0.0018
ethnicity_simple  14.833  5 0.0111
GLOBAL           172.669 12 <2e-16
```

#### Schoenfeld residuals

##### Non-avoidable ED attendances

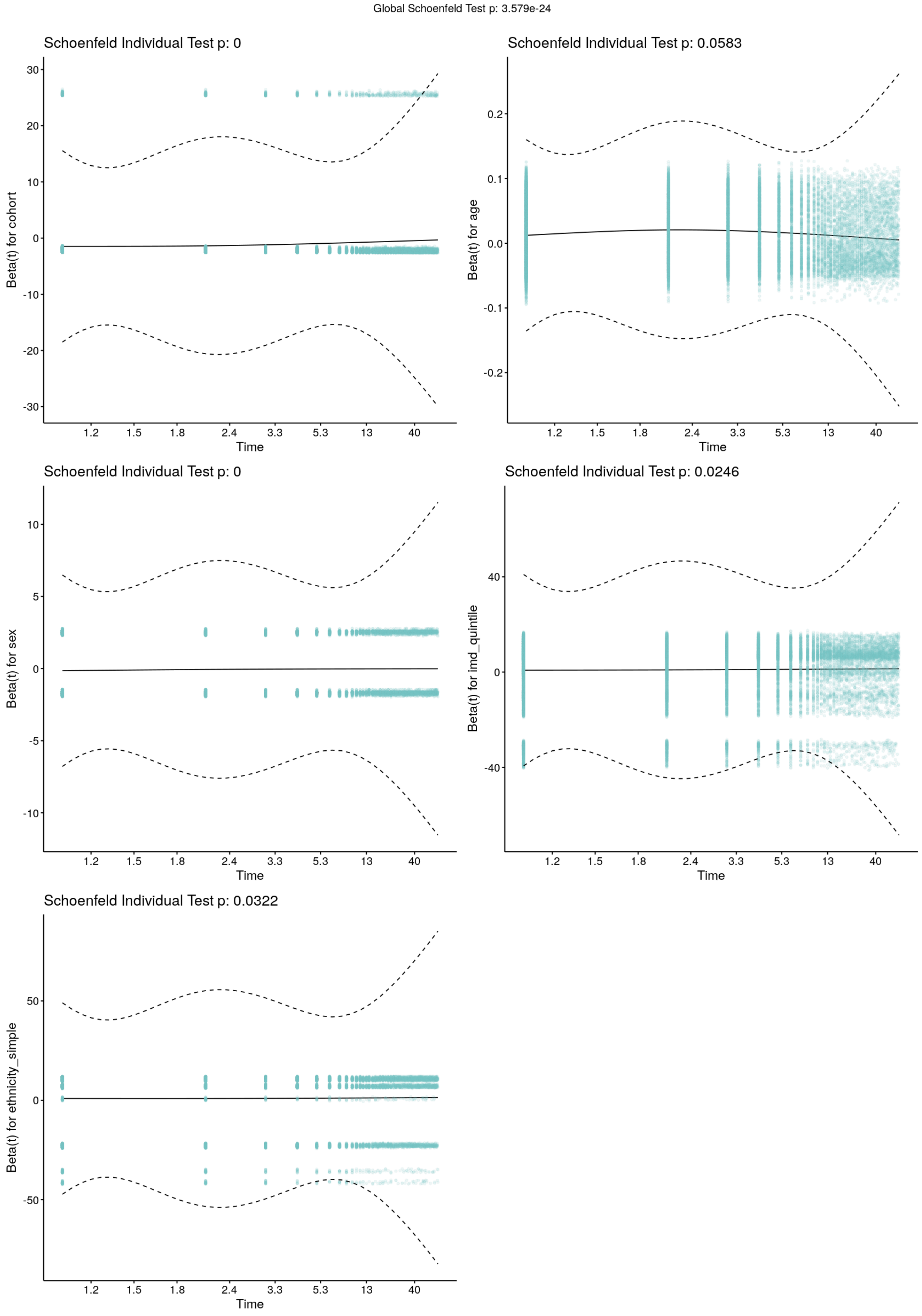


### All ED attendances

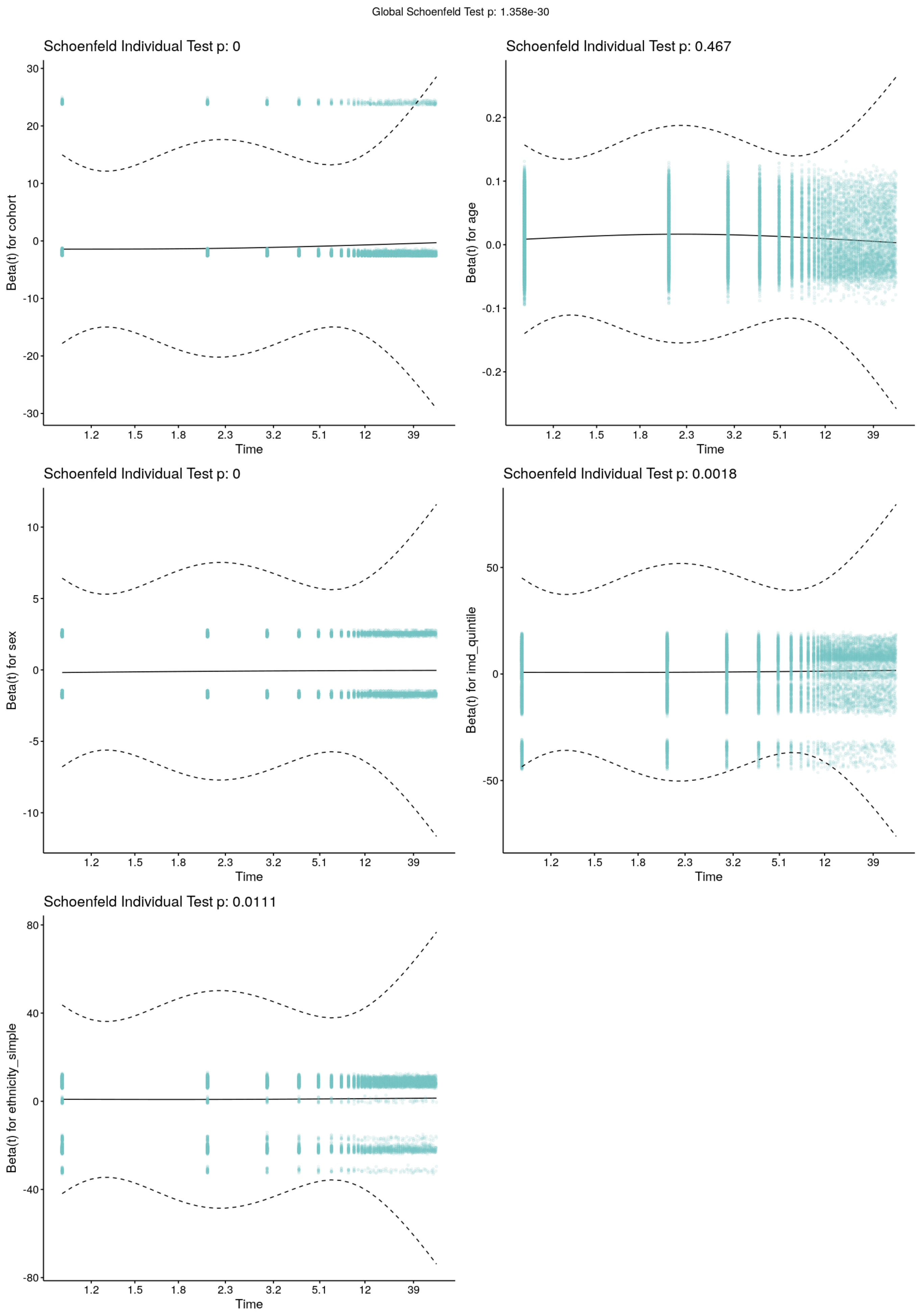

Global Schoenfeld Test p: 1.358e-30
Schoenfeld Individual Test p: 0
Beta(t) for cohort
30
20
10
0
-10
-20
-30
1.2
1.5
1.8
2.3
3.2
5.1
12
39
Time
Schoenfeld Individual Test p: 0.467
Beta(t) for age
0.2
0.1
0.0
-0.1
-0.2
1.2
1.5
1.8
2.3
3.2
5.1
12
39
Time
Schoenfeld Individual Test p: 0
Beta(t) for sex
10
5
0
-5
-10
1.2
1.5
1.8
2.3
3.2
5.1
12
39
Time
Schoenfeld Individual Test p: 0.0018
Beta(t) for imd_quintile
50
0
-50
1.2
1.5
1.8
2.3
3.2
5.1
12
39
Time
Schoenfeld Individual Test p: 0.0111
Beta(t) for ethnicity_simple
80
40
0
-40
-80
1.2
1.5
1.8
2.3
3.2
5.1
12
39
Time
